# A Comprehensive Investigation of Machine Learning Practices in Predictive Modeling of Alzheimer’s Disease and Related Dementias using Multisite Real-world Electronic Health Records

**DOI:** 10.64898/2025.11.30.25341312

**Authors:** Qiannan Zhang, Zhenxing Xu, Weishen Pan, Hiroko H. Dodge, Jiayu Zhou, Chang Su, Fei Wang

## Abstract

The irreversible progression and profound societal impact of Alzheimer’s disease and related dementias (AD/ADRD) underscore the pressing need for early risk prediction. Machine learning (ML) models integrated with electronic health records (EHRs) offer promising solutions for generating real-world evidence, yet their clinical utility for AD/ADRD remains unclear, both in terms of whether they can be reliably applied and how they can be broadly integrated into healthcare practice. In this study, we systematically investigated widely used ML models for AD/ADRD early prediction using the nationwide All of Us EHRs as the primary cohort, exploring multiple analytic dimensions including cohort construction, feature engineering strategies, and subtype-specific modeling, spanning prediction windows ranging from 10 years to 1 day. To assess clinical applicability in healthcare practice as well as provide actionable insights for cross-cohort model adaptation, we transferred pretrained models from All of Us to external EHR repositories (INSIGHT CRN and OHSU RDW) using different transfer paradigms. Collectively, the findings delineate the strengths and limitations of EHR-based ML models, demonstrating their capability for near-term prediction and interpretability while revealing constraints in long-term estimation, and offer empirical insights that support practical model use with multisite EHRs.

## Introduction

Alzheimer’s Disease and Alzheimer’s Disease-related Dementias (AD/ADRD) currently affect more than 55 million individuals worldwide^1^. In the United States, as of 2025, over 7.2 million adults aged 65 years and older (approximately 11% of this age group) are living with AD/ADRD, while the annual incidence is projected to double by 2050 as the population ages^2^. Individuals with AD/ADRD typically experience progressive cognitive and functional deterioration, accompanied by behavioral and psychological symptoms, culminating in loss of independence and a substantial need for long-term care. It results in major emotional, physical, and financial burdens on patients and caregivers, posing an escalating public health challenge^3^.

Against this backdrop, early prediction of AD/ADRD is crucial. Identifying high-risk individuals during the preclinical or prodromal stages, when neuropathological alterations may remain reversible or modifiable, enables timely lifestyle or therapeutic interventions and supports informed care planning for patients and families^4^. Early risk estimation and stratification also facilitate more effective disease-modifying clinical trials through the identification of appropriate study populations^5^. At the population level, such prediction informs resource allocation and policy development aimed at mitigating the long-term societal and economic impacts of dementia. As such, considerable efforts have been dedicated to identifying predictive biomarkers, including protein signatures ^6–9^ and neuroimaging markers^10–12^. While these approaches show promise, their clinical utility in real-world settings remains limited, constrained by challenges such as insufficient longitudinal follow-up and restricted data availability^13^.

The widespread adoption of Electronic Health Records (EHR) systems has enabled access to real-world patient data (RWD) encompassing disease status, comorbidities, medications, laboratory results, and treatment outcomes. Collected during routine care, these data provide a scalable and cost-efficient foundation for generating real-world evidence (RWE)^14,15^ and advancing research on complex diseases such as AD/ADRD^16,17^. Concurrently, the rise of machine learning (ML) has introduced powerful analytic tools capable of uncovering patterns and associations in healthcare data that often elude conventional statistical methods^18^. Recent ML-based models leveraging EHR data have shown potential in early identification of individuals at risk for AD/ADRD^19–21^. Despite these advances, variations in cohort selection, feature representation, model design, and evaluation protocols have led to divergent predictive performance and risk factor profiles. Although frameworks such as the OMOP Common Data Model (CDM)^22^ have aimed to facilitate data harmonization, model transferability across cohorts is still underexplored, despite its importance for clinical applicability. Key uncertainties persist around their clinical utility, in terms of whether they can be reliably applied and how they can be broadly integrated in healthcare practice.

To fill these gaps, the present study systematically evaluated ML practices for developing predictive models of AD/ADRD using multisite EHRs (Fig. 1), including major AD/ADRD subtypes, namely Alzheimer’s disease (AD), vascular dementia (VaD), Lewy body dementia (LBD), frontotemporal dementia (FTD), and mixed-type dementia (multiple subtypes). Specifically, we leveraged three large-scale, real-world EHR repositories that capture diverse demographic and clinical populations across the United States: the nationwide All of Us research program^23^; the INSIGHT Clinical Research Network (INSIGHT CRN)^24^, comprising over 15 million patients from five major academic medical centers in New York City; and the Oregon Health & Science University Research Data Warehouse (OHSU RDW)^25^, representing a regional academic medical system. We used All of Us as the cohort for model development and internal evaluation, capitalizing on its broad representativeness and detailed longitudinal data. To characterize model reliability across both short- and long-term horizons, we applied prediction windows from 10 years to 1 day and examined how feature engineering and cohort construction choices affect performance. We additionally developed subtype-specific models to account for the clinical heterogeneity of AD/ADRD^20^. To explore their broader applicability in healthcare practice, pretrained models were evaluated on the two external cohorts using different transfer strategies alongside de novo retraining. Finally, we analyzed feature importance patterns across prediction windows, modeling setups, and cohorts to examine the consistency and robustness of identified predictors.

**Fig. 1.**
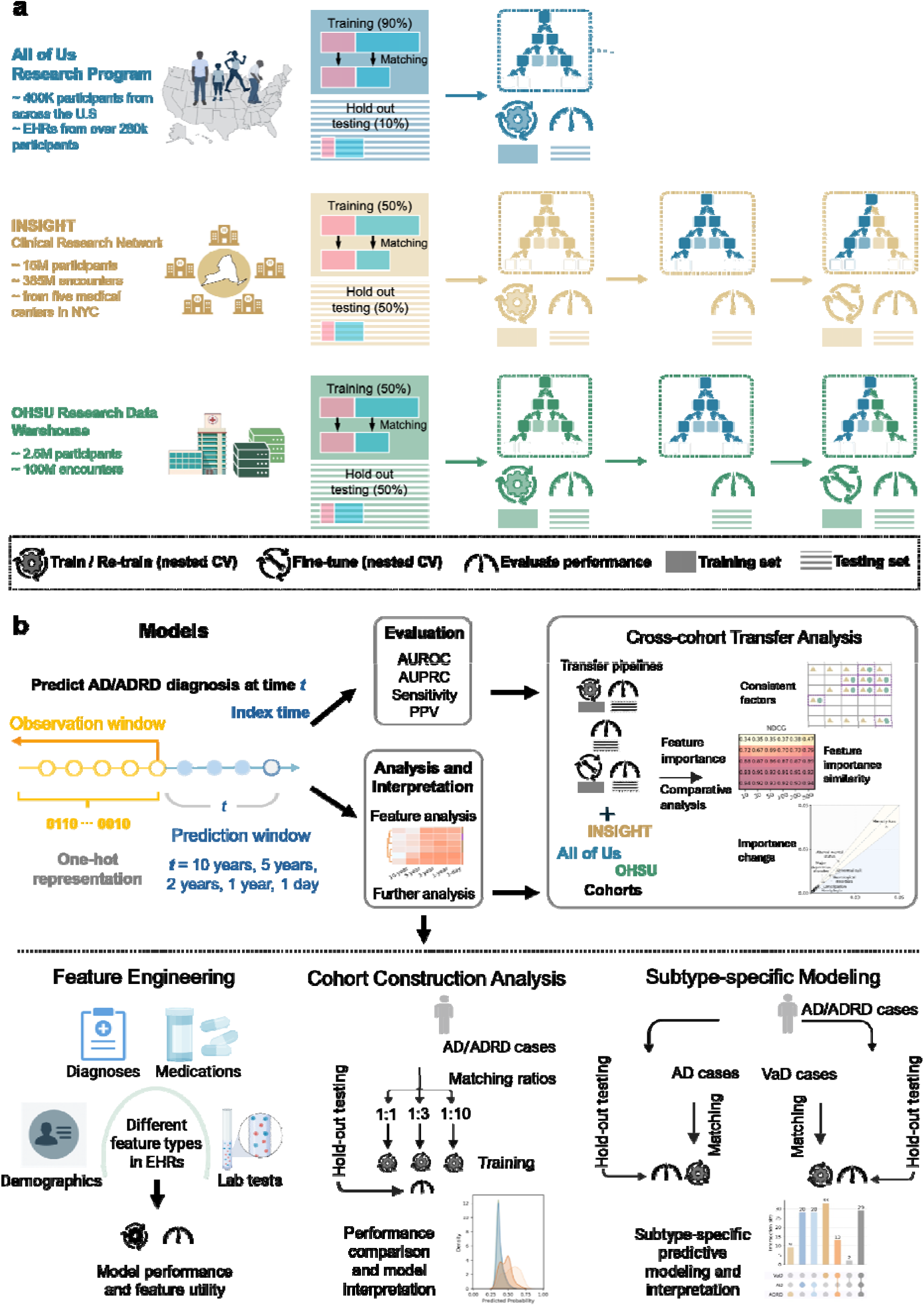
Overview of study design. (**a**) Electronic health record (EHR) data from the All of Us Research Program, the INSIGHT Clinical Research Network, and the OHSU Research Data Warehouse were used to develop and evaluate ML models for predicting the onset of AD/ADRD. Within each cohort, matched training sets and randomly held-out test sets were constructed, and model development employed 5-fold nested cross-validation for each prediction window. To examine cross-cohort transferability, three paradigms were applied to INSIGHT and OHSU: direct transfer, which applied pretrained models without modification; fine-tuning, which adapted pretrained models using external training data; and retraining, which rebuilt models solely from external matched training sets. (**b**) Prediction tasks were defined across multiple prediction windows (10 years, 5 years, 2 years, 1 year, and 1 day) using one-hot encoded patient features extracted from corresponding observation windows. Model performance was assessed using multiple metrics, with further analyses of model design choices, including feature type contributions, cohort construction strategies, and subtype-specific discriminability for AD and vascular dementia (VaD). Cross-cohort transfer analysis further investigated model applicability and provided actionable insights for model use with multisite EHRs. Feature importance was analyzed across prediction windows, subtypes, and cohorts, aimed to reveal how ML models capture predictors across populations, and meanwhile resolve the differences that varied by prediction windows and subtypes.

Collectively, our analysis demonstrated that ML models delivered reliable near-term prediction, while being limited by sparse and subtle signals as the window extended (10 to 5 years). By dissecting feature and cohort choices, we characterized how modeling designs shaped model behavior and predictive performance. Cross-cohort evaluations further clarified the trade-offs in adapting pretrained models to external health systems: direct transfer could suffice for short windows or data-limited sites, whereas fine-tuning or retraining offered advantages for earlier windows with richer local data. Feature analyses revealed that ML models captured predictors robust across populations, while also resolving meaningful differences that varied by prediction window and subtype. Taken together, these findings delineate the current capability boundaries of EHR-based ML models: providing actionable near-term risk estimates, transferring across sites with modest adaptation, and identifying interpretable, biologically plausible predictors.

## Results

### Study cohorts

From the three repositories, we constructed study cohorts spanning diverse U.S. populations. Using established AD/ADRD definitions^20^ (code list in Supplementary Table 7), we identified an initial pool of 1,436 AD/ADRD cases in All of Us, defined as participants with at least one AD/ADRD diagnosis and aged 50 years or older at the identified index time, and 234,033 individuals as the initial control pool. Controls had no AD/ADRD diagnoses, no dementia-related conditions (e.g., mild cognitive impairment [MCI], Parkinson’s disease), and no exposure to anti-dementia medications. We randomly held out 10% of the initial pools for evaluation on All of Us, and used the remainder to construct matched training sets for each prediction window, pairing each case with up to 10 controls without replacement using propensity scores^26–29^ and controlling for demographics (birth year, sex, race, and ethnicity) and Charlson comorbidity index^30,31^ (excluding dementia-related conditions^20^).

Consistent inclusion and exclusion criteria were applied to the INSIGHT and OHSU repositories, yielding initial pools of 35,621 cases and 2,517,393 controls in INSIGHT, and 3,315 cases and 122,416 controls in OHSU. In both cohorts, 50% of cases and controls were randomly reserved for hold-out testing, and the remainder underwent propensity score matching to create matched training sets (Methods). Summary statistics are provided in Table 1, full cohort characteristics in Supplementary Table 2, and inclusion/exclusion flows in Extended Data Fig. 1.

**Table 1.** Demographic summaries for individuals used in models, and the example matched cohorts for the 1-day prediction window. OHSU contains limited demographic variables. See full cohort description in Supplement Table 2. SMD = standardized mean difference

| Individuals after initial filtering |  | All of Us |  | INSIGHT |  |  | OHSU |  |  |
| --- | --- | --- | --- | --- | --- | --- | --- | --- | --- |
|  | Control | Case |  | Control | Case |  | Control | Case |  |
| *n* | 234033 | 1436 |  | 2517393 | 35621 |  | 122416 | 3315 |  |
| *Birth year, mean (SD)* | 1967.6 (16.7) | 1946.0 (9.2) |  | 1960.9 (13.6) | 1939.6 (9.7) |  | - | - |  |
| *Age of onset, mean (SD)* | - | 71.8 (9.8) |  | - | 79.7 (9.7) |  | - | 79.3 (8.7) |  |
| **Sex, *n* (%)** |  |  |  |  |  |  |  |  |  |
| *Female* | 143613 (61.4) | 698 (48.6) |  | 1452086 (57.7) | 21677 (60.9) |  | - | - |  |
| *Male* | 85778 (36.7) | 702 (48.9) |  | 1064263 (42.3) | 13934 (39.1) |  | - | - |  |
| *Other/unknown* | 4642 (2.0) | 36 (2.5) |  | 1044 (0.0) | 10 (0.0) |  | - | - |  |
| **Ethnicity, *n* (%)** |  |  |  |  |  |  |  |  |  |
| *Hispanic or Latino* | 44789 (19.1) | 234 (16.3) |  | 312630 (12.4) | 7693 (21.6) |  | - | - |  |
| *Not Hispanic or Latino* | 180502 (77.1) | 1148 (79.9) |  | 1220005 (48.5) | 18380 (51.6) |  | - | - |  |
| *Other/unknown* | 8742 (3.7) | 54 (3.8) |  | 984758 (39.1) | 9548 (26.8) |  | - | - |  |
| **Race, *n* (%)** |  |  |  |  |  |  |  |  |  |
| *Black* | 45237 (19.3) | 230 (16.0) |  | 263441 (10.5) | 4825 (13.5) |  | - | - |  |
| *Asian* | 6468 (2.8) | 25 (1.7) |  | 2681 (0.1) | 31 (0.1) |  | - | - |  |
| *Middle Eastern/North African* | 1363 (0.6) | 5 (0.3) |  | - | - |  | - | - |  |
| *Other/unknown* | 52683 (22.5) | 292 (20.3) |  | 1152793 (45.8) | 15182 (42.6) |  | - | - |  |
| *White* | 128282 (54.8) | 884 (61.6) |  | 1098478 (43.6) | 15583 (43.7) |  | - | - |  |
| Matched cohorts (1-day) |  | All of Us |  |  | INSIGHT |  |  | OHSU |  |
|  | Control | Case | SMD | Control | Case | SMD | Control | Case | SMD |
| *n* | 11537 | 1157 |  | 100957 | 11841 |  |  |  |  |
| *Birth year, mean (SD)* | 1945.8 (9.1) | 1945.8 (9.2) | -0.004 | 1941.3 (8.5) | 1939.4 (9.3) | -0.215 | - | - | - |
| *Index time/onset age, mean (SD)* | 72.3 (9.7) | 72.3 (9.8) | 0.003 | 78.2 (8.5) | 80.1 (9.4) | 0.213 | 78.9 (8.6) | 79.1 (8.6) | 0.023 |
| **Sex, *n* (%)** |  |  |  |  |  |  |  |  |  |
| *Female* | 5866 (50.8) | 556 (48.1) | -0.056 | 57911 (57.4) | 7222 (61.0) | 0.074 | - | - | - |
| *Male* | 5554 (48.1) | 580 (50.1) | 0.040 | 43029 (42.6) | 4616 (39.0) | -0.074 | - | - | - |
| *Other/unknown* | 117 (1.0) | 21 (1.8) | 0.068 | 17 (0.0) | 3 (0.0) | 0.006 | - | - | - |
| **Ethnicity, *n* (%)** |  |  |  |  |  |  |  |  |  |
| *Hispanic or Latino* | 1150 (10.0) | 183 (15.8) | 0.175 | 16661 (16.5) | 2782 (23.5) | 0.175 | - | - | - |
| *Not Hispanic or Latino* | 9864 (85.5) | 931 (80.4) | -0.134 | 54440 (53.9) | 6060 (51.2) | -0.055 | - | - | - |
| *Other/unknown* | 523 (4.5) | 43 (3.7) | -0.041 | 29856 (29.6) | 2999 (25.3) | -0.095 | - | - | - |
| **Race, *n* (%)** |  |  |  |  |  |  |  |  |  |
| *Black* | 1981 (17.2) | 198 (17.1) | -0.002 | 12780 (12.7) | 1715 (14.5) | 0.053 | - | - | - |
| *Asian* | 132 (1.1) | 19 (1.6) | 0.043 | 81 (0.1) | 15 (0.1) | 0.014 | - | - | - |
| *Middle Eastern/North African* | 11 (0.1) | 3 (0.3) | 0.039 | - | - | - | - | - | - |
| *Other/unknown* | 1708 (14.8) | 229 (19.8) | 0.132 | 39838 (39.5) | 4941 (41.7) | 0.046 | - | - | - |
| *White* | 7705 (66.8) | 708 (61.2) | -0.117 | 48258 (47.8) | 5170 (43.7) | -0.083 | - | - | - |

We defined an index time to identify features preceding the earliest clinical indication. For cases, the index time was the earliest encounter with an AD/ADRD diagnosis or an anti-dementia medication prescription. Matched controls were assigned the encounter date closest to the case’s index time, whereas hold-out controls used the date one year before their last EHR encounter to support clinical relevance and model development (see details in Methods). Prediction windows (10 years, 5 years, 2 years, 1 year, and 1 day) specified how far in advance the model made predictions. Thus, observation windows extended from the first recorded EHR encounter to the start of each window, and individuals with under one year of observation were excluded. EHR variables in the observation window were converted into categorized, one-hot encoded inputs, with deliberated feature preprocessing methods applied (Methods). Models were developed using nested 5-fold cross-validation (CV)^32^ on matched All of Us training sets, evaluated on the outer loops and hold-out test sets, with the design choices systematically assessed (Extended Data Fig. 2). We applied the best-performing models to hold-out test sets in INSIGHT and OHSU for direct transfer, and for fine-tuning using each cohort’s matched training set. As a comparator, de novo models were trained using only external cohort’s data. Evaluation metric included area under the receiver operating characteristic curve (AUROC), area under the precision-recall curve (AUPRC), and sensitivity and positive predictive values (PPV) at fixed specificities (90% and 95%).

### ML models predict short-term and long-term risk of AD/ADRD

Three ML models were employed, including one linear model (i.e., Logistic Regression, LR) and two non-linear models, Random Forest (RF) and XGBoost. On the All of Us hold-out test set, RF models achieved AUROCs ranging from 0.665±0.016 at the 10-year prediction window to 0.879±0.003 at the 1-day window (Fig. 2). The corresponding AUPRCs were 0.026±0.002 and 0.187±0.009, exceeding the reference AD/ADRD prevalence of 0.013 and 0.011 for the 10-year and 1-day windows. At 90% specificity, RF models yielded sensitivities of 0.184±0.026 (10-year) and 0.691±0.009 (1-day), while the values changed to 0.124±0.023 and 0.535±0.014 at 95% specificity. PPV values were 0.027±0.004 and 0.071±0.001 at 90% specificity, with further increases at 95%. LR models underperformed relative to RF, especially at earlier prediction windows, reflecting the limited capacity of linear models to capture complex EHR feature interactions. XGBoost performed comparably to RF models, particularly for shorter prediction windows (1 year and 1 day). Full metrics, including outer-loop validation results, are reported in Supplementary Table 1.

**Fig. 2.**
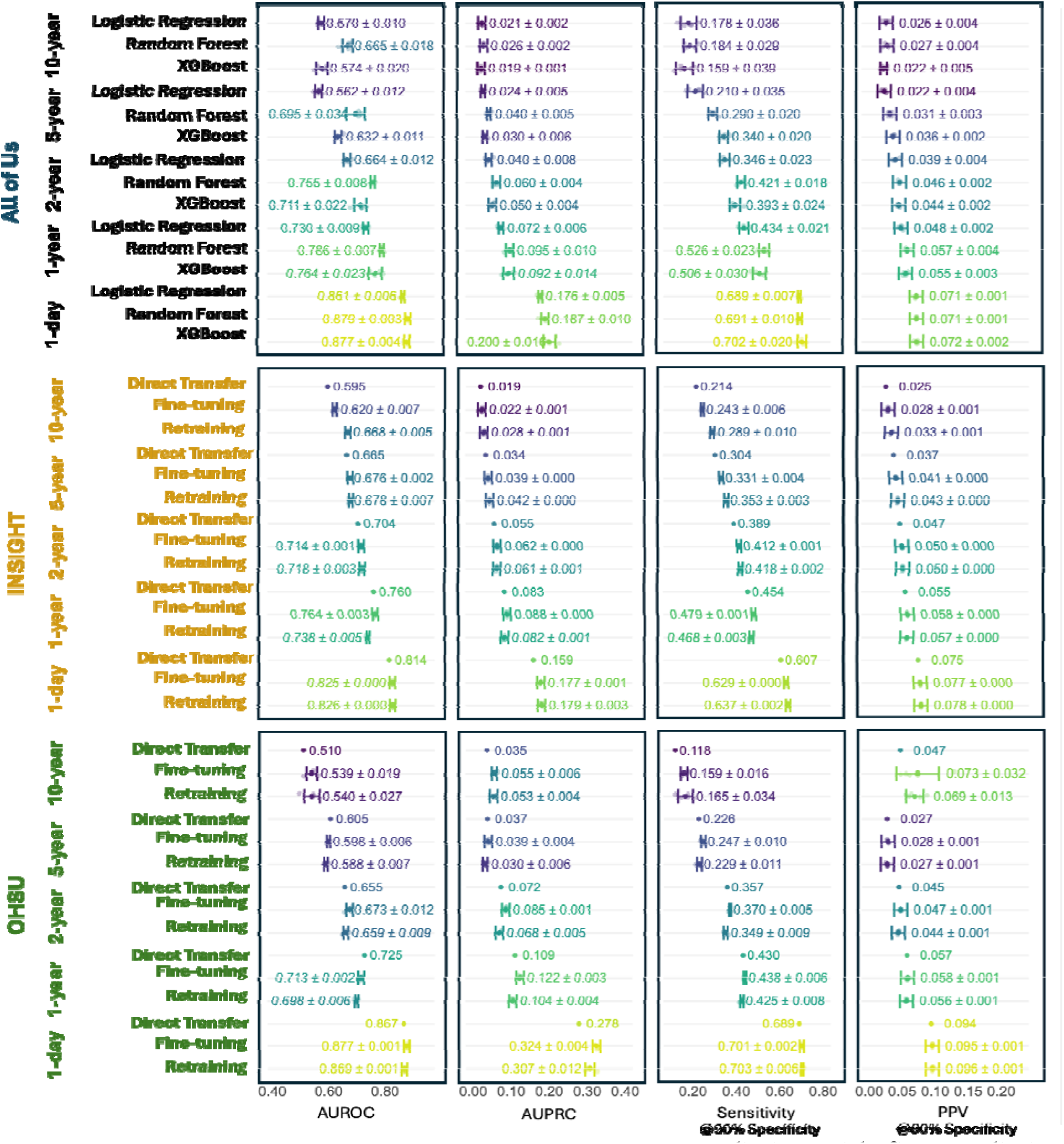
The performance of ML models for AD/ADRD prediction with five prediction windows across three cohorts. Models trained on each outer fold of the 5-fold nested cross-validation were evaluated on the hold-out test sets. Average performance and standard deviation (SD) across folds are reported, with individual fold results as scatter points. Linear and nonlinear ML models were assessed in the All of Us cohort, and different learning paradigms, including direct transfer and fine-tuning of the pretrained best-performing Random Forest models in All of Us, as well as de novo retraining for comparison, were further evaluated on the external cohort (Methods). Supplementary Tables 1, 5, and 6 provide the full results, including per-fold performance on the hold-out test sets and the outer-fold validation sets.

Leveraging the matched cohort to mitigate potential confounding, we examined predictive factors for AD/ADRD onset, using feature importance scores from RF models (Methods). Given their direct clinical relevance and predictive utility (Extended Data Fig. 6), diagnostic features were analyzed at the Phecode-category level^33^, and temporal patterns of top-ranked Phecodes were characterized via hierarchical clustering (Methods). As shown in Fig. 3a, Phecode groups related to mental, circulatory, and metabolic disorders emerged as the most predictive of AD/ADRD across prediction windows. Within the top 50 specific Phecodes, one cluster consistently exhibited strong predictive capability, comprising memory loss, depression, type 2 diabetes (T2D), dizziness and giddiness, and coronary atherosclerosis (Fig. 3b). Certain conditions showed relative importance with specific prediction windows. For instance, dysthymic disorder, migraine, tobacco use disorder, convulsions, peripheral neuropathy, and bipolar disorder had a greater impact 10 to 5 years before AD/ADRD onset. In contrast, obstructive sleep apnea, urinary incontinence, and altered mental status gained prominence at shorter windows. Besides, another cluster maintained moderate importance across windows, such as joint pain, obesity, hyperlipidemia, osteoarthrosis, and back pain.

**Fig. 3.**
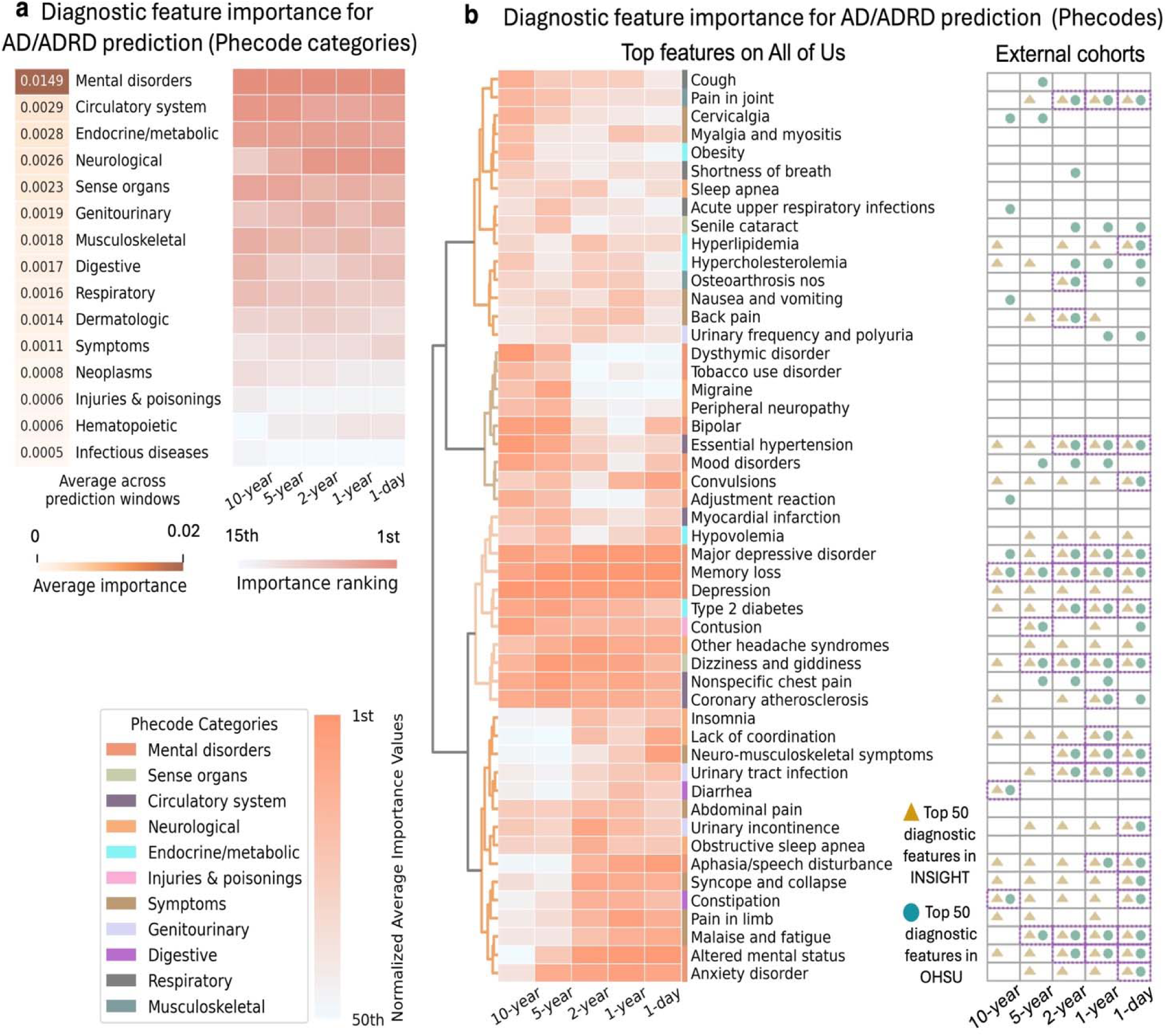
ML model trained on matched All of Us data facilitated hypothesis generation for AD/ADRD predictors. (**a**) Top Phecode categories sorted by their average importance across prediction windows (Methods). (**b**) (Left) Top 50 specific Phecodes identified across prediction windows in All of Us, using hierarchical clustering with Ward linkage on feature importance rankings; (Right) External replication in INSIGHT (yellow triangles) and OHSU (blue circles), indicating whether each feature appeared among the top 50 predictors of each external cohort. Features consistently ranked in the top 50 across datasets (highlighted in dashed purple boxes) represent robust and generalizable predictive signals.

### Diagnosis and medication prescription records are crucial predictors

As EHRs contain diverse data types during routine care, assessing their relative contributions is essential for interpretable and effective AD/ADRD prediction. To this end, we compared the predictive value of diagnoses, medications, laboratory tests, and demographics (Fig. 4a; Methods). Diagnosis-based RF models achieved an AUROC of 0.614±0.034 and AUPRC of 0.021±0.003 at the 10-year window (reference prevalence: 0.013), increasing to 0.873±0.002 and 0.129±0.004 for 1-day models (reference prevalence: 0.011). Medication-based RF models showed lower performance, with AUROCs of 0.594±0.023 and 0.699±0.006 and AUPRCs of 0.021±0.002 and 0.049±0.004 at the 10-year and 1-day windows, respectively. In contrast, RF models trained solely with laboratory tests or demographic features demonstrated limited predictive capability. Similar trends were observed for LR and XGBoost models, with full results provided in Supplementary Table 1.

**Fig. 4.**
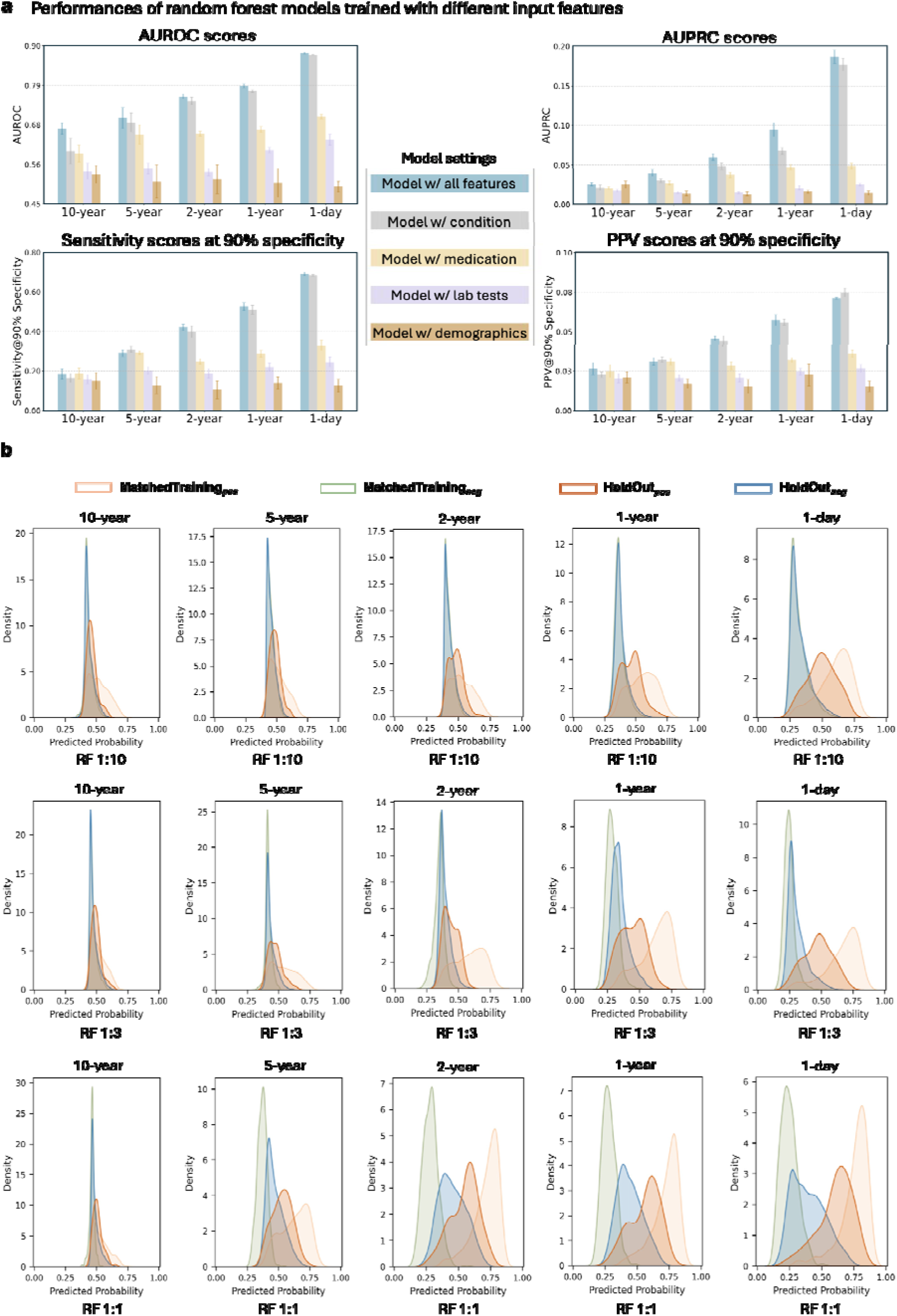
Analysis of feature types and cohort construction strategies. **(a)** Performance comparison between full RF models and feature type-specific RF models on the All of Us hold-out test set. Bars show the average and standard deviation (SD) across models from the 5 outer folds of the nested cross-validation. **(b)** Predicted probability from RF models trained with different case–control matching ratios (1:1, 1:3, and 1:10) across prediction windows. Distributions are displayed using kernel density estimation for case (positive) and control (negative) samples scored by the best-performing outer-fold model on its own training splits (MatchedTraining) and on the hold-out test set (HoldOut).

As prior literature frequently relied on well-established AD/ADRD-associated features^19,20,34,35^, we excluded their Phecodes from model inputs to quantify their specific contribution and whether predictive signal remains beyond these features (code list in Supplementary Table 8). This exclusion substantially degraded RF model performances, e.g., AUROCs dropping from 0.879±0.003 to 0.774±0.002 and AUPRCs from 0.187±0.009 to 0.060±0.005 for the 1-day window. Comparable declines were observed for LR and XGBoost models (Supplementary Table 1). Yet, the performance reduction was less pronounced for longer prediction windows (10 to 5 years), which may be explained by the rising prevalence of these codes closer to disease onset (Extended Data Fig. 3). These findings suggest that well-established diagnostic features turn predictive near disease onset, primarily signaling late-stage rather than long-term risk.

### Matching ratios modulate model overfitting behaviors

As matching ratio affects both confounding control and sample size, we assess its impact on model behaviors by comparing RF models under 1:1 and 1:3 alternatives, in addition to the baseline 1:10 ratio. RF models demonstrated robust performance across ratios, with AUROCs and AUPRCs remaining broadly comparable (Supplementary Table 4). We further visualized predicted probabilities via kernel density estimation using RF models that yielded the best hold-out performance (Fig. 4b; Methods). Overall, the predicted-probability distributions for cases and controls (denoted as positive and negative classes) in the hold-out test set underscored the intrinsic difficulty of early AD/ADRD prediction. With longer prediction windows, case and control distributions became more concentrated and increasingly overlapping. The distributional patterns were also influenced by matching ratios. Under 1:1 matching, predicted scores for cases and controls were well separated in the training set but overlapped in testing, suggesting distribution shifts and signs of overfitting. By contrast, higher ratios (1:3 and 1:10) produced better alignment between training and testing distributions for controls, indicating improved generalization. At the same time, case scores tended to decrease, bringing their distribution closer to that of the controls. These trends likely reflect a training process more consistent with the underlying data distribution, yielding a relatively realistic decision boundary and avoiding overconfident separation during model fitting. Taken together, the results suggest that matching ratios play a role in modulating overfitting behaviors in AD/ADRD prediction models.

### ML models reveal subtype-specific predictive patterns of AD and VaD

In view of AD/ADRD heterogeneity, we further investigated the discriminative ability of ML models across subtypes, focusing on AD and VaD^36–42^, and constructed subtype-specific predictive models (see cohort details in Supplementary Table 2; Methods). AD-specific RF models produced AUROCs ranging from 0.608±0.045 (10-year) to 0.836±0.011 (1-day), with corresponding AUPRCs of 0.009±0.001 (Reference AD prevalence: 0.006) and 0.027±0.001 (Reference AD prevalence: 0.005). VaD-specific RF models exhibited better performance than AD prediction, reaching AUROCs of 0.760±0.089 and 0.876±0.006 at the 10-year and 1-day windows, respectively, while AUPRCs varied from 0.017±0.003 (Reference VaD prevalence: 0.006) to 0.112±0.004 (Reference VaD prevalence: 0.006). LR and XGBoost showed similar performance trends relative to RF (Fig. 5a-b; Supplementary Table 4).

**Fig. 5.**
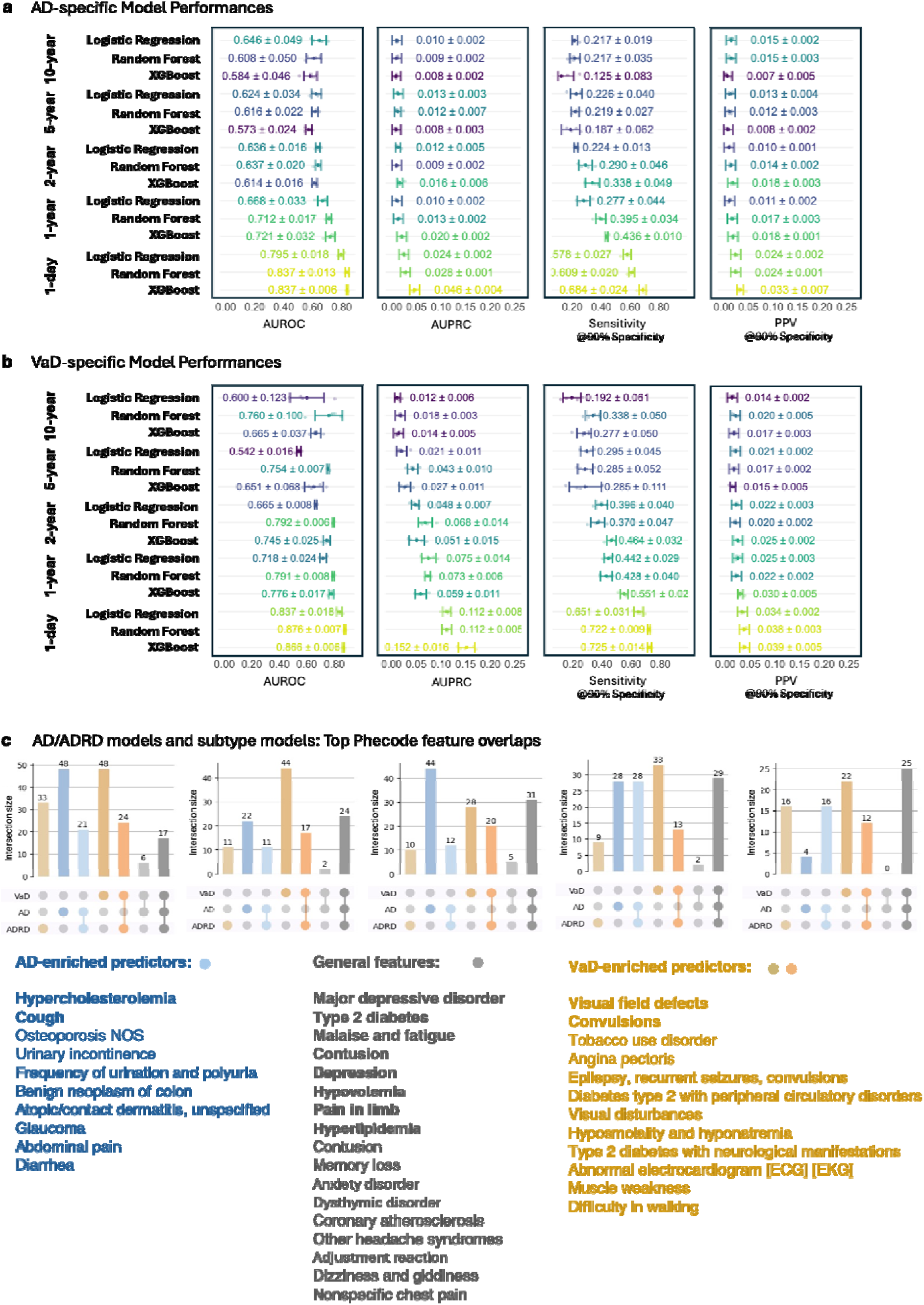
Analysis of predictive modeling for AD/ADRD subtypes. (**a-b**) Performance of AD-specific and vascular dementia (VaD)-specific models across five prediction windows. Error bars represent standard deviation (SD) across outer folds of nested cross-validation, with scatter points indicating performance of the models from individual folds; (**c**) Overlap of important features identified by AD/ADRD models and subtype-specific models (Methods). Important features were selected from the best-performing outer-fold model as those whose importances exceeded the 95th-percentile threshold at each prediction window. The UpSet plot summarizes the overlap. Shared and subtype-enriched predictors are listed alongside, with **boldface** indicating appearance across all five windows and regular font indicating consistency in four.

Using subtype-specific RF models, we examined the importance of Phecode categories and feature temporal patterns (Extended Data Fig. 4–5; Supplementary Table 3). Across both AD and VaD, mental and metabolic disorders emerged as principal contributors, with memory loss, depression, and T2D remaining consistent individual predictors across prediction windows. In AD-specific models, musculoskeletal, genitourinary, circulatory, and dermatologic categories showed higher importance in earlier windows, whereas digestive and neurological problems gained importance closer to onset. Temporal analyses at the Phecode level revealed predictive features for early systemic dysfunctions (e.g., inflammatory musculoskeletal, autonomic, and neuropsychiatric abnormalities such as fasciitis and erectile dysfunction), chronic physiological burden (e.g., peripheral neuropathy, osteoarthrosis, hypothyroidism), and symptoms that increased in importance near disease onset (e.g., diarrhea, abdominal pain, altered mental status). By contrast, VaD models emphasized circulatory and neurological categories, with coronary atherosclerosis, myocardial infarction, and essential hypertension as leading predictors. Temporal clusters for VaD noted late-stage features (e.g., anxiety, lack of coordination) along with predictors detectable up to 10 years before index time (e.g., extrapyramidal syndrome, precordial pain, diabetic polyneuropathy). Additional moderately influential features comprised visual disturbances, tobacco use disorder, and other vascular diseases.

In further comparison with general AD/ADRD models (Fig. 5c; Supplementary Table 3), shared predictors across all models included depression, T2D, hypovolemia, and memory loss. AD-enriched features (either shared by the AD and AD/ADRD models or specific to the AD models) encompassed factors related to metabolic, genitourinary and degenerative processes, such as hypercholesterolemia, osteoporosis, urinary incontinence, urinary frequency/polyuria. In contrast, VaD-enriched features (e.g., visual field defects, tobacco use disorder, abnormal electrocardiogram, difficulty in walking) predominantly reflected vascular and circulatory impairment^43–45^. These findings indicate that ML models capture subtype-specific signatures, such as neurodegenerative and vascular patterns.

### Pretrained models show strong transferability for short-term prediction but limited utility for earlier windows

To evaluate cross-cohort generalizability, the best-performing models were selected from All of Us and applied to INSIGHT and OHSU (Methods). Without adaptation, the transferred model on INSIGHT yielded AUROCs ranging from 0.595 (AUPRC: 0.019, reference prevalence: 0.011) to 0.814 (AUPRC: 0.159, reference prevalence: 0.013) as prediction windows varied from 10 years to 1 day (Fig.2, Supplementary Table 5). De novo retraining mostly outperformed direct transfer in AUROC and AUPRC, especially showing notable gain at the 10-year window (AUROCs: 0.668±0.004; AUPRCs: 0.028±0.001). Fine-tuning led to performance improvements relative to direct transfer, producing results comparable to retraining.

For OHSU, direct transfer of RF models achieved AUROCs of 0.510 (AUPRC: 0.035; reference prevalence: 0.024) and 0.867 (AUPRC: 0.278; reference prevalence: 0.015) for 10-year and 1-day windows, respectively, performing on par with or better than retrained models in shorter prediction windows (5 years to 1 day). At the 10-year window, however, direct transfer proved less effective, whereas retraining produced slightly higher performance (AUROCs: 0.540±0.024; AUPRCs: 0.053±0.004). Fine-tuning improved performance over direct transfer across prediction windows and generally surpassed retraining when predicting within 5 years. Detailed results are provided in Supplementary Table 6.

To investigate how features contribute across cohorts and paradigms, feature importance was explored by comparing the best-performing retrained models with their pretrained counterparts, and with newly generated trees from fine-tuned models (Methods), focusing on Phecodes given their predictive capability across cohorts (Extended Data Fig. 6, Supplementary Table 5-6). Between pretrained and retrained models, a portion of top predictors in All of Us stayed highly ranked in external cohorts, such as memory loss, dizziness and giddiness, etc. (Fig. 3b). Quantitatively, as the prediction window lengthened, feature similarity measured by set overlap (overlap similarity, weighted Jaccard index) and ranking agreement (NDCG) declined (Fig. 6a-b) declined. For instance, the top-k features (k = {10, 30, 50, 100, 200, 500}) of pretrained and retrained models overlapped by 53%–76% on INSIGHT and 44–70% on OHSU for the 1-day window, while dropping to 20%-44% and 0-50% for the 10-year window, respectively. At the individual-feature level (Fig. 6c-d), retrained models elevated the importance of abnormal gait while reducing others (e.g., major depressive disorder). It also revealed cohort-specific predictors, such as oncologic screening and nutrition-related symptoms, reflecting variation in clinical documentation and coding practices across health systems. Compared with retraining, fine-tuning introduced new trees that largely preserved the predictive features identified by retrained models, across both quantitative and feature-level analysis, though the feature similarity diminished at longer windows, suggesting greater reliance on pretrained knowledge as local long-term data became sparser.

**Fig. 6.**
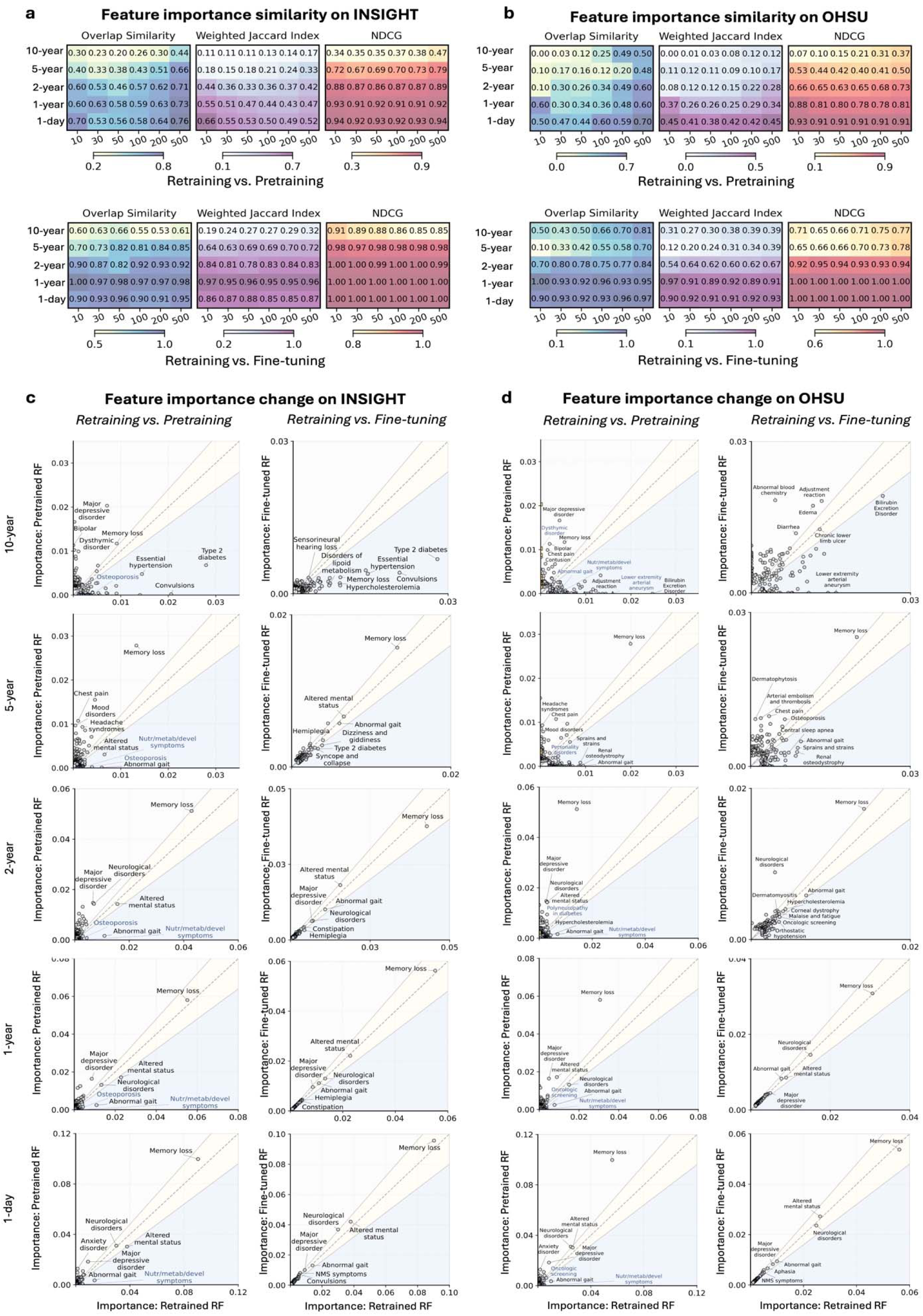
Feature importance analysis in cross-cohort model transfer. (**a-b**) Quantitative comparison of feature importance similarity between retrained and pretrained models, as well as between retrained and fine-tuning paradigms on external cohorts. Feature importance was derived from the pretrained model for direct transfer, and from the best-performing outer-fold model for fine-tuning and retraining within each external cohort. The heatmaps show set-level feature similarity, measured by overlap similarity and weighted Jaccard index, and feature ranking agreement (NDCG) across top-k features (k = 10–500) from each paradigm. **(c-d)** The scatter plots compare relative feature importance for overlapped features between direct transfer, fine-tuning, and retraining paradigms, with diagonal reference lines indicating agreement.

## Discussion

Real-world data, particularly EHRs, offer a practical foundation for building early AD/ADRD risk prediction. Despite their potential, the feasibility and limitations of developing such models from EHRs have not been systematically characterized. In this study, we addressed this gap by building and analyzing AD/ADRD prediction models with the primary All of Us cohort and two external repositories (INSIHGT and OHSU). Across all datasets, models performed substantially better near disease onset, with marked degradation over longer prediction windows (10 to 5 years). This consistent trend underscores the intrinsic challenges in long-term risk estimation, likely driven by subtle prodromal signals, delayed or incomplete clinical assessments, and reduced healthcare utilization years before diagnosis.

After being trained on All of Us, we explored such models’ transferability, which is critical for model applicability across heterogeneous health systems. Through evaluation under three paradigms, we aim to provide experimental evidence and actionable insights to guide the practical model reuse in local circumstances. Our results suggest that at the longer window (e.g., 10 years), pretrained models, whether fine-tuned or not, showed poor transferability and were outperformed by retraining on both external cohorts, reflecting widening cohort differences over time (Fig. 2). For shorter-term prediction (within 5 years), direct transfer performed comparably to retraining in data-limited settings such as OHSU, where the small local sample size may be insufficient to support effective training from scratch. In contrast, retraining produced relative gains in data-rich cohorts like INSIGHT. In either scenario, fine-tuning tended to outperform direct transfer, though the extent of its benefit over retraining varied across cohorts and prediction windows. Building on these observations, we provide empirically grounded guideline for selecting transfer strategies for model use across cohorts, emphasizing prediction window, data availability, and computational resources as key determinants (Extended Data Fig. 7). At the feature level, a greater proportion of factors predictive in All of Us were preserved in INSIGHT than in OHSU across the 10- to 1-year windows (Fig. 3b), which was as well indicated by quantitative analyses (Fig. 5). This likely reflects INSIGHT’s multi-institutional and demographically diverse population, which more closely parallels the nationwide composition of All of Us, in contrast to the single-center profile of OHSU. These results highlight that geographic, demographic, and practice-related differences manifest as shifts between cohorts, in turn influencing cross-cohort model transfer.

After adjusting for confounders, our model identified AD/ADRD predictors consistent with prior findings, such as diabetes mellitus and memory loss^46,47^. It also revealed bipolar disorder, associated with elevated dementia risk (6.6% vs. 4% in controls over a 13-year follow-up^48^), and depression, which has been independently linked to AD/ADRD incidence^49^. The identified early manifestations (e.g., migraine, tobacco use disorder)^50–53^ and late-stage symptoms (e.g., lack of coordination, diarrhea)^54,55^ also aligned with existing literature. Furthermore, the subtype analyses uncovered discriminative factors in alignment with current research. For instance, myocardial infarction showed an association with VaD (e.g., adjusted HR: 1.35) but had less consistent links to AD^56^. Midlife hypertension was a shared risk factor for both subtypes, yet more strongly tied to VaD due to its circulatory basis^57,58^. In contrast, our identification of hypercholesterolemia as an AD-enriched factor aligned with evidence associating it with amyloid plaque formation and AD onset^59–61^. Osteoporosis, another AD-enriched factor, shares inflammatory and bone–brain metabolic dysregulation pathways with AD^19,62,63^. These findings demonstrate the capacity of ML models to discover clinically meaningful and biologically plausible signals for AD/ADRD.

Our analysis also showed diagnoses as the dominant predictors, followed by medications, whereas laboratory tests and demographics contributed little. Unlike the targeted clinical information in diagnoses and medications, laboratory tests were generally nonspecific. It points to the need for modality-aware modeling over naive feature fusion. We also found that matching ratios in cohort construction may modulate overfitting behaviors. Although small ratios more strictly reduce confounding, they also narrow the sample space, potentially suppressing data diversity. Conversely, higher ratios mitigate this issue but demand attention to class imbalance during training.

This study has several limitations. First, inherent issues in EHR data, such as missingness and variability, may introduce bias in prediction. Second, the analysis is based on structured EHRs, potentially overlooking signals from clinical notes, imaging, genetics, and biomarkers. Also, the work focused on widely applied linear and non-linear ML models, rather than deep neural networks, given their potential deficiency on high-dimensional, sparse EHR data^64^. Finally, the external datasets may not fully represent geographic and institutional diversities of broader healthcare systems.

Overall, our study provides a rigorous, large-scale, multisite assessment of ML models for AD/ADRD prediction using real-world EHR data. By systematically assessing factors that influence model performance and clinical utility, we identify key determinants of model effectiveness and transferability across healthcare systems. These findings deepen understanding of how ML models perform with heterogeneous EHR data. While current models can generate actionable near-term risk estimates, adapt across sites, and highlight interpretable, biologically plausible predictors, their long-term predictive power remains constrained by EHR sparsity and model inexpressiveness. Future research can explore more expressive learning architectures and complementary modalities to strengthen long-term prediction. Together, this work establishes a foundation for developing robust, interpretable, and clinically meaningful ML tools for AD/ADRD early prediction. By validating clinical utility, this work advances translational AI for aging and precision health research.

## Methods

### Patient identification

The study utilizes EHR data primarily from the All of Us Research Program, a flagship U.S. precision medicine initiative launched by the National Institutes of Health (NIH) in 2018. For more than 400k enrolled participants in the Controlled Tier dataset (version 7), the program integrates various data modalities, such as EHRs, surveys, and genetics. Approximately half of these participants have contributed EHR data encompassing a broad range of demographic and clinical attributes, including medical conditions, medications, vital signs, and laboratory measurements. The All of Us program adopts the Observational Medical Outcomes Partnership (OMOP) Common Data Model (CDM) for data standardization.

Our study targets early prediction of AD/ADRD, covering leading subtypes including Alzheimer’s Disease (AD), Vascular Dementia (VaD), Lewy Body Dementia (LBD), Frontotemporal Dementia (FTD), and mixed-type dementia (multiple subtypes). Prior studies have typically defined AD/ADRD cases utilizing diagnostic codes or the prescribing of anti-dementia medications, although their scope often varied. Some investigations focused exclusively on AD^19,65^, whereas others considered a broader spectrum of dementia types, such as alcohol-related dementia^66–69^. We utilized diagnostic codes to identify AD/ADRD cases corresponding to the aforementioned specific AD/ADRD subtypes while avoiding inclusion of less related dementia types by carefully selecting established computable phenotypes^20,70^ (see code list for case and control definition in Supplementary Table 7).

Specifically, individuals with at least one AD/ADRD diagnosis were included in the initial case pool. The initial control pool comprised individuals with no AD/ADRD diagnosis, no other conditions associated with or causal to dementia (e.g., MCI, Parkinson’s disease, alcoholism, etc.), and no exposure to anti-dementia medications. For both All of Us and each external repository, we randomly reserve a fixed proportion from the initial case and control pools to form the hold-out test set. The remaining samples are used to construct the matched training set for model development. Since All of Us serves as our primary cohort, we retain 90% of the data for matched cohort construction and allocate the remaining 10% for hold-out testing.

### Observation and prediction window

We define the case’s index time as the earlier of the first recorded AD/ADRD diagnosis or the first prescription of an anti-dementia drug. Given the age-related nature of AD/ADRD, cases were required to be at least 50 years old at the index time. To construct the matched cohort, each case was matched with controls at a 1:10 ratio, balancing for birth year, gender, race and ethnicity, and the Charlson Comorbidity Index (CCI), with dementia-related conditions excluded to minimize bias. Propensity scores were estimated using logistic regression, and controls were selected as nearest neighbors to the corresponding case without replacement. In the matched cohort, controls were required to be within one year of their corresponding case’s age and to have at least one medical encounter within six months of the case’s index time. Each control’s index time was then assigned as the encounter date closest to that of its matched case, ensuring temporal and demographic comparability between cases and controls^20^. For the hold-out test set, the control’s index time was defined as one year before their last recorded visit^19^, reflecting a pragmatic design compatible with diverse healthcare systems. Controls should also satisfy the condition of an index age of 50 years or above. This definition of control index time aimed for clinical relevance while maintaining the integrity and reliability of the ML models. We employed varying prediction windows of 10 years, 5 years, 2 years, 1 year, and 1 day before index time to examine the effects of temporal proximity on prediction. Observation windows spanned from each individual’s first EHR visit up to the start of the prediction window, during which clinical data were collected for both cases and controls. Inclusion and exclusion flows, and cohort characteristics are provided for all three datasets (Extended Data Fig. 1, Supplementary Table 2).

### Data extraction and preparation

We utilized all variables within the observation window with appropriate preprocessing for predictive modeling. Demographic variables, e.g., gender, race, and ethnicity, were categorized using one-hot encoding. Age at prediction, defined as the participant’s age at model application, was normalized to the range of 0 to 1. To mitigate data sparsity, clinical data were aggregated into higher-level concepts through a code grouping strategy consistent with previous studies^20,71^. Specifically, standard OMOP condition concepts in AoU were mapped to SNOMED codes, then converted to ICD-10 codes using rule-based mappings from the National Library of Medicine (NLM) (https://www.nlm.nih.gov/healthit/snomedct/us_edition.html; September 2024 release). The resulting ICD-10 codes were subsequently mapped to PheWas (Phenome-wide association studies) codes^33,72–74^. Medication data were harmonized by mapping OMOP drug concepts to RxNorm clinical and branded drug identifiers, which were further consolidated to ingredient-level RXCUIs using the NLM RxNav APIs^75^. For continuous variables, the most recent real-valued measurements within each observation window, including vital signs (e.g., BMI, systolic and diastolic blood pressures) and routine laboratory tests, were categorized as normal, abnormally high, or abnormally low according to reference ranges^76^ (see Supplementary Table 9 for laboratory tests and reference ranges). All categorical features were further transformed into a binary format through one-hot encoding. Clinical features that were missing or fell within normal ranges were encoded as 0 to indicate default values. Finally, to ensure sufficient longitudinal coverage, participants in both the matched cohort and hold-out test set had to have at least one year of EHR data in the observation window (at least two visits separated by a minimum of one year).

### ML model preparation and training

To predict AD/ADRD onset, we trained binary classification models via nested cross-validation (CV) on the matched cohort at each prediction window, employing five folds in both the inner and outer loops. The outer loops of nested CV divided the matched cohort into five stratified folds: four folds were used for training and hyperparameter tuning via an inner 5-fold cross-validation, and the remaining fold served for validation. The model from each outer-loop fold was subsequently evaluated on the hold-out test set to provide an unbiased performance estimate.

Three machine learning models, namely Logistic Regression, Random Forest, and XGBoost, were trained to predict AD/ADRD onset using scikit-learn and xgboost packages, with balanced class weights applied where appropriate. Hyperparameters were optimized via grid search, maximizing AUROC during cross-validation. For tree-based models, grid search parameters were designed to account for overfitting and feature correlation, including n_estimators (0.5 * feature_num, feature_num, 2 * feature_num), max_depth (ranging from 3 to 30 in increments of 7), and max_features (sqrt, log_2_) that adjusts the subset used in each tree. Logistic Regression and XGBoost were further tuned for learning rate (0.001, 0.005, 0.01) to optimize convergence. Model performance was evaluated using AUROC, AUPRC, and sensitivity and PPV at specificity thresholds of 90% and 95%. Results are reported for all five outer-loop folds as well as the hold-out test set to provide a comprehensive evaluation of model performance.

### Experimental setup variants

Experimental settings varied to comprehensively evaluate the capability of ML models, considering the following aspects.

#### Feature specification

To assess the impact of different input feature types, we trained ML models separately using only demographic, condition, medication, or laboratory test features on the matched cohorts, while keeping the training and testing data split unchanged. In addition, models were retrained after excluding a predefined set of explicit Phecode features (Supplementary Table 8) and evaluated using the same procedure.

#### Matching Ratio variation

We examined the impact of different matching ratios on model performance by repeating case–control matching within the same 90% split of the initial pools at each prediction window using alternative ratios of 1:3 and 1:1 to construct model development cohorts. With each prediction window, RF models were retrained on each matched cohort, and the best-performing model on the hold-out test set was selected to visualize predicted probability distributions using kernel density estimation^77^. We visualized that distribution for training samples from the corresponding outer-loop and testing samples from the hold-out set to illustrate model behaviors during training and testing.

#### Subtype stratification

To assess the model’s ability to distinguish specific disease subtypes, ML models were developed for AD and VaD, respectively. From the same 90% split of the initial case and control pools, patients with a specific subtype were re-matched to controls at a 1:10 ratio to create subtype-specific matched cohorts at each prediction window. The resulting models were evaluated on the hold-out test set to distinguish subtype cases from controls.

### Preparation for external repositories

#### Cohort construction

INSIGHT Clinical Research Network (CRN)^78^, established in 2014 and funded by the Patient-Centered Outcomes Research Institute (PCORI)^79^, is a multi-institutional research network aggregating EHRs from five academic medical centers located in New York City. The network includes approximately 15 million patient records and more than 385 million clinical encounters, conforming to the PCORnet CDM. We applied a pipeline consistent with that used for All of Us for cohort construction and data preprocessing. Specifically, 35,621 cases and 2,517,393 controls were initially identified, with 50% of participants randomly held out for testing. The remaining participants underwent 1:10 propensity score matching based on demographics and CCI scores to form a matched cohort. The same criteria as in AoU, including index time, age filtering, and EHR length requirement, were applied. Demographic and clinical variables were processed using strategies aligned with those of AoU and prepared as model input for the five prediction windows.

The Oregon Health & Science University (OHSU) Research Data Warehouse, curated by the Oregon Clinical and Translational Research Institute (OCTRI), contains longitudinal EHR data for more than 2.5 million individuals. From OHSU data, 3315 cases and 123,648 controls were identified in the initial pools, followed by a 50/50 random split for cohort construction. Due to the limited availability of demographic and measurement data, only age was consistently present, necessitating propensity score matching based solely on age and CCI scores. Besides, as medical encounters in OHSU are recorded using age-based timestamps, the index time for controls was defined as the age corresponding to the visit nearest to the case’s index age, with a difference of no more than one year. Model inputs were restricted to condition and medication data. Aside from these, all other criteria followed the same protocol as used for INSIGHT.

#### ML model preparation

We applied the same feature preprocessing pipeline to external cohorts where applicable. Model optimization on external datasets was performed using nested 5-fold cross-validation. For direct transfer, the best-performing RF model at each prediction window from All of Us was directly tested on external hold-out test set for evaluation after harmonizing external cohort input features into the pretrained model. In the fine-tuning and de novo retraining paradigms, RF models were either fine-tuned or retrained through nested CV using the external matched cohorts, then evaluated on the external hold-out test set. Specifically for fine-tuning, additional trees were trained on the external data and incorporated into the pretrained RF ensemble by majority voting, with performance assessed both during CV and on the external test set. Hyperparameters were determined via grid search in n_estimators (0.5 * feature_num, feature_num, 2 * feature_num), max_depth (3 to 20, step size = 5) and max_feature (sqrt, log_2_) for fine-tuning and retraining paradigms, respectively.

### Feature interpretation

We employed RF models for their interpretability and capacity to capture non-linear relationships. Feature importance was quantified as the average decrease in Gini impurity across decision tree splits, reflecting each feature’s contribution to model decisions. We focused primarily on diagnostic (Phecode) features, which directly reflect disease progression and risk than medications or laboratory measurements, which often represent downstream consequences of a diagnosis.

#### Category-level aggregation

To capture broader clinical relevance, feature importance was first aggregated at the Phecode category level. For each prediction window, we averaged the importance of the top five Phecodes within each category to derive a category-level score. These were then ranked by their mean importance across windows to highlight clinically meaningful domains contributing to AD/ADRD or subtype-specific predictions.

#### Temporal analysis

Feature importance was further analyzed at the individual Phecode level to examine temporal patterns. For each prediction window, the top 50 Phecodes, ranked by their mean importance across time windows, were identified. Hierarchical clustering with Ward linkage was then applied to the ranking trajectories across prediction windows to uncover structural relationships and temporal groupings among features.

#### Subtype-Driven Signatures

To assess feature consistency and specificity across disease subtypes, we extracted Phecodes whose importance exceeded the 95th percentile in each RF model that performed the best on the hold-out test set at each time window for AD/ADRD, AD, and VaD prediction. Overlaps were visualized via UpSet plots to identify shared and subtype-specific predictors. A feature was considered *subtype-enriched* if it was important in a subtype model or the general AD/ADRD model, while being absent or less significant for the other subtype models.

#### Cross-Dataset Comparison

To explore feature robustness under dataset shifts, feature importance was compared under different learning paradigms in INSIGHT and OHSU. For each prediction window, the top 50 diagnostic predictors from the best-performing retrained model in each external dataset were compared against the top 50 from the original All of Us model, to identify shared predictors across cohorts (Fig. 3b). Broader comparisons (ranks 10–500) were conducted using multiple similarity metrics (Fig. 6a-b): Overlap similarity is applied to measure the co-existence of features, weighted Jaccard Index further introduces importance scores to weight different features, while NDCG directly measures ranking correlation. Furthermore, shared features were visualized using scatter plots to directly compare importance scores across models (Fig. 6c-d). Likewise, Retrained models were also compared with fine-tuning generated trees to explore how data and learning paradigms interact.

## Data Availability

Individual-level data from the All of Us Research Program are available to approved researchers whose institutions have established data use agreements with the program. Access to INSIGHT and OHSU data will require approved data use agreement and may be directed to the INSIGHT and OHSU Institutional Review Board.

## Reference

1. Focus on Alzheimer’s Disease and Related Dementias | National Institute of Neurological Disorders and Stroke. https://www.ninds.nih.gov/current-research/focus-disorders/focus-alzheimers-disease-and-related-dementias.

2. Hebert, L. E., Beckett, L. A., Scherr, P. A. & Evans, D. A. Annual incidence of Alzheimer disease in the United States projected to the years 2000 through 2050. Alzheimer Dis. Assoc. Disord. 15, 169–173 (2001).

3. Alzheimer’s Association 2025 Alzheimer’s Disease Facts and Figures.

4. Diogo, V. S., Ferreira, H. A., Prata, D., & for the Alzheimer’s Disease Neuroimaging Initiative. Early diagnosis of Alzheimer’s disease using machine learning: a multi-diagnostic, generalizable approach. Alzheimers Res. Ther. 14, 107 (2022).

5. de Aquino, C. H. Methodological Issues in Randomized Clinical Trials for Prodromal Alzheimer’s and Parkinson’s Disease. Front. Neurol. 12, 694329 (2021).

6. Winchester, L. M. et al. Artificial intelligence for biomarker discovery in Alzheimer’s disease and dementia. Alzheimers Dement. 19, 5860–5871 (2023).

7. Chang, C.-H., Lin, C.-H. & Lane, H.-Y. Machine Learning and Novel Biomarkers for the Diagnosis of Alzheimer’s Disease. Int. J. Mol. Sci. 22, 2761 (2021).

8. Stamate, D. et al. A metabolite-based machine learning approach to diagnose Alzheimer-type dementia in blood: Results from the European Medical Information Framework for Alzheimer disease biomarker discovery cohort. Alzheimers Dement. Transl. Res. Clin. Interv. 5, 933–938 (2019).

9. Cedazo-Minguez, A. & Winblad, B. Biomarkers for Alzheimer’s disease and other forms of dementia: Clinical needs, limitations and future aspects. Exp. Gerontol. 45, 5–14 (2010).

10. Arbabshirani, M. R., Plis, S., Sui, J. & Calhoun, V. D. Single subject prediction of brain disorders in neuroimaging: Promises and pitfalls. NeuroImage 145, 137–165 (2017).

11. Pellegrini, E. et al. Machine learning of neuroimaging for assisted diagnosis of cognitive impairment and dementia: A systematic review. Alzheimers Dement. Diagn. Assess. Dis. Monit. 10, 519–535 (2018).

12. Mueller, S. G. et al. Ways toward an early diagnosis in Alzheimer’s disease: The Alzheimer’s Disease Neuroimaging Initiative (ADNI). Alzheimers Dement. 1, 55–66 (2005).

13. Drzezga, A. & Barthel, H. Imaging and Fluid Biomarkers of Alzheimer Disease: Complementation Rather Than Competition. J. Nucl. Med. Off. Publ. Soc. Nucl. Med. 66, S32–S44 (2025).

14. Concato, J. & Corrigan-Curay, J. Real-World Evidence — Where Are We Now? N. Engl. J. Med. 386, 1680–1682 (2022).

15. Sherman, R. E. et al. Real-World Evidence — What Is It and What Can It Tell Us? N. Engl. J. Med. 375, 2293–2297 (2016).

16. Hampel, H. et al. The impact of real-world evidence in implementing and optimizing Alzheimer’s disease care. Med N. Y. N 6, 100695 (2025).

17. Galvin, J. E. et al. Generating real-world evidence in Alzheimer’s disease: Considerations for establishing a core dataset. Alzheimers Dement. 20, 4331–4341 (2024).

18. Beam, A. L. & Kohane, I. S. Big Data and Machine Learning in Health Care. JAMA 319, 1317–1318 (2018).

19. Tang, A. S. et al. Leveraging electronic health records and knowledge networks for Alzheimer’s disease prediction and sex-specific biological insights. Nat. Aging 4, 379–395 (2024).

20. Li, Q. et al. Early prediction of Alzheimer’s disease and related dementias using real-world electronic health records. Alzheimers Dement. 19, 3506–3518 (2023).

21. Akter, S., Liu, Z., Simoes, E. J. & Rao, P. Using machine learning and electronic health record (EHR) data for the early prediction of Alzheimer’s Disease and Related Dementias. J. Prev. Alzheimers Dis. 12, 100169 (2025).

22. Reinecke, I. et al. The Usage of OHDSI OMOP – A Scoping Review. in German Medical Data Sciences 2021: Digital Medicine: Recognize – Understand – Heal 95–103 (IOS Press, 2021). doi:10.3233/SHTI210546.

23. Ramirez, A. H. et al. The All of Us Research Program: Data quality, utility, and diversity. Patterns 3, (2022).

24. Min, J. et al. Association between antidiabetic drug use and the risk of COVID-19 hospitalization in the INSIGHT Clinical Research Network in New York City. Diabetes Obes. Metab. 24, 1402–1405 (2022).

25. Oregon Clinical and Translational Research Institute (OCTRI). (2023). Research Data Warehouse (RDW) [Data set]. Oregon Health and Science University.

26. Choi, B. Y. Propensity score analysis for health care disparities: a deweighting approach. BMC Med. Res. Methodol. 24, 106 (2024).

27. Granger, E., Watkins, T., Sergeant, J. C. & Lunt, M. A review of the use of propensity score diagnostics in papers published in high-ranking medical journals. BMC Med. Res. Methodol. 20, 132 (2020).

28. Yao, X. I., et al. Reporting and Guidelines in Propensity Score Analysis: A Systematic Review of Cancer and Cancer Surgical Studies. JNCI J. Natl. Cancer Inst. 109, djw323 (2017).

29. Rosenbaum, P. R. & Rubin, D. B. The central role of the propensity score in observational studies for causal effects. Biometrika 70, 41–55 (1983).

30. Charlson, M. E., Carrozzino, D., Guidi, J. & Patierno, C. Charlson Comorbidity Index: A Critical Review of Clinimetric Properties. Psychother. Psychosom. 91, 8–35 (2022).

31. Glasheen, W. P. et al. Charlson Comorbidity Index: ICD-9 Update and ICD-10 Translation. Am. Health Drug Benefits 12, 188–197 (2019).

32. Parvandeh, S., Yeh, H.-W., Paulus, M. P. & McKinney, B. A. Consensus features nested cross-validation. Bioinformatics 36, 3093–3098 (2020).

33. Bastarache, L. Using Phecodes for Research with the Electronic Health Record: From PheWAS to PheRS. Annu. Rev. Biomed. Data Sci. 4, 1–19 (2021).

34. Tang, A. et al. Leveraging Electronic Medical Records and Knowledge Networks to Predict Disease Onset and Gain Biological Insight Into Alzheimer’s Disease. 2023.03.14.23287224 Preprint at 10.1101/2023.03.14.23287224 (2023).

35. Nianogo, R. A. et al. Risk Factors Associated With Alzheimer Disease and Related Dementias by Sex and Race and Ethnicity in the US. JAMA Neurol. 79, 584 (2022).

36. Bir, S. C., Khan, M. W., Javalkar, V., Toledo, E. G. & Kelley, R. E. Emerging Concepts in Vascular Dementia: A Review. J. Stroke Cerebrovasc. Dis. 30, (2021).

37. Iadecola, C. The Pathobiology of Vascular Dementia. Neuron 80, 844–866 (2013).

38. O’Brien, J. T. & Thomas, A. Vascular dementia. The Lancet 386, 1698–1706 (2015).

39. Kalaria, R. N. et al. Alzheimer’s disease and vascular dementia in developing countries: prevalence, management, and risk factors. Lancet Neurol. 7, 812–826 (2008).

40. Rizzi, L., Rosset, I. & Roriz-Cruz, M. Global Epidemiology of Dementia: Alzheimer’s and Vascular Types. BioMed Res. Int. 2014, 908915 (2014).

41. Román, G. C. Vascular dementia may be the most common form of dementia in the elderly. J. Neurol. Sci. 203–204, 7–10 (2002).

42. Román, G. C. Vascular Dementia: Distinguishing Characteristics, Treatment, and Prevention. J. Am. Geriatr. Soc. 51, S296–S304 (2003).

43. Lary, C. W. et al. Bone Mineral Density and the Risk of Incident Dementia: a meta-analysis. J. Am. Geriatr. Soc. 72, 194–200 (2024).

44. Xiao, T. et al. Association of Bone Mineral Density and Dementia: The Rotterdam Study. Neurology 100, e2125–e2133 (2023).

45. Thavorn, K. et al. Upper Gastrointestinal Bleeding in Elderly Adults with Dementia Receiving Cholinesterase Inhibitors: A Population-Based Cohort Study. J. Am. Geriatr. Soc. 62, 382–384 (2014).

46. Tiwari, S. S. et al. Alzheimer-related decrease in CYFIP2 links amyloid production to tau hyperphosphorylation and memory loss. Brain 139, 2751–2765 (2016).

47. Sims-Robinson, C., Kim, B., Rosko, A. & Feldman, E. L. How does diabetes accelerate Alzheimer disease pathology? Nat. Rev. Neurol. 6, 551–559 (2010).

48. Nielsen, J. L., Kaltoft, K., Wium-Andersen, I. K., Wium-Andersen, M. K. & Osler, M. Association of early- and late-life bipolar disorder with incident dementia. A Danish cohort study. J. Affect. Disord. 367, 367–373 (2024).

49. Elser, H., et al. Association of Early-, Middle-, and Late-Life Depression With Incident Dementia in a Danish Cohort. JAMA Neurol. 80, 949–958 (2023).

50. Hurh, K. et al. Increased risk of all-cause, Alzheimer’s, and vascular dementia in adults with migraine in Korea: a population-based cohort study. J. Headache Pain 23, 108 (2022).

51. Kim, J., Ha, W. S., Park, S. H., Han, K. & Baek, M. S. Association between migraine and Alzheimer’s disease: a nationwide cohort study. Front. Aging Neurosci. 15, 1196185 (2023).

52. Rusanen, M., Kivipelto, M., Quesenberry, C. P., Jr, Zhou, J. & Whitmer, R. A. Heavy Smoking in Midlife and Long-term Risk of Alzheimer Disease and Vascular Dementia. Arch. Intern. Med. 171, 333–339 (2011).

53. Ott, A. et al. Smoking and risk of dementia and Alzheimer’s disease in a population-based cohort study: the Rotterdam Study. The Lancet 351, 1840–1843 (1998).

54. Collyer, T. A. et al. Association of Dual Decline in Cognition and Gait Speed With Risk of Dementia in Older Adults. JAMA Netw. Open 5, e2214647 (2022).

55. Kowalski, K. & Mulak, A. Brain-Gut-Microbiota Axis in Alzheimer’s Disease. J. Neurogastroenterol. Motil. 25, 48–60 (2019).

56. Sundbøll, J. et al. Higher Risk of Vascular Dementia in Myocardial Infarction Survivors. Circulation 137, 567–577 (2018).

57. Launer, L. J. et al. Midlife blood pressure and dementia: the Honolulu-Asia aging study. Neurobiol. Aging 21, 49–55 (2000).

58. Qiu, C., Winblad, B. & Fratiglioni, L. The age-dependent relation of blood pressure to cognitive function and dementia. Lancet Neurol. 4, 487–499 (2005).

59. Pappolla, M. A., Refolo, L., Sambamurti, K., Zambon, D. & Duff, K. Hypercholesterolemia and Alzheimer’s Disease: Unraveling the Connection and Assessing the Efficacy of Lipid-Lowering Therapies. J. Alzheimers Dis. JAD 101, S371–S393 (2024).

60. Feringa, F. M. & van der Kant, R. Cholesterol and Alzheimer’s Disease; From Risk Genes to Pathological Effects. Front. Aging Neurosci. 13, 690372 (2021).

61. Wang, H. et al. Regulation of beta-amyloid production in neurons by astrocyte-derived cholesterol. Proc. Natl. Acad. Sci. 118, e2102191118 (2021).

62. Dengler-Crish, C. M. & Elefteriou, F. Shared mechanisms: osteoporosis and Alzheimer’s disease? Aging 11, 1317–1318 (2019).

63. Wanionok, N. E., Morel, G. R. & Fernández, J. M. Osteoporosis and Alzheimeŕs disease (or Alzheimeŕs disease and Osteoporosis). Ageing Res. Rev. 99, 102408 (2024).

64. Xiao, C., Choi, E. & Sun, J. Opportunities and challenges in developing deep learning models using electronic health records data: a systematic review. J. Am. Med. Inform. Assoc. JAMIA 25, 1419–1428 (2018).

65. Park, J. H. et al. Machine learning prediction of incidence of Alzheimer’s disease using large-scale administrative health data. Npj Digit. Med. 3, 1–7 (2020).

66. Nori, V. S. et al. Machine learning models to predict onset of dementia: A label learning approach. Alzheimers Dement. Transl. Res. Clin. Interv. 5, 918–925 (2019).

67. Ben Miled, Z., et al. Predicting dementia with routine care EMR data. Artif. Intell. Med. 102, 101771 (2020).

68. Albrecht, J. S., Hanna, M., Kim, D. & Perfetto, E. M. Predicting Diagnosis of Alzheimer’s Disease and Related Dementias Using Administrative Claims. J. Manag. Care Spec. Pharm. 24, 1138–1145 (2018).

69. Matthews, K. A. et al. Racial and ethnic estimates of Alzheimer’s disease and related dementias in the United States (2015–2060) in adults aged ≥65 years. Alzheimers Dement. 15, 17–24 (2019).

70. Wilkinson, T. et al. Identifying dementia outcomes in UK Biobank: a validation study of primary care, hospital admissions and mortality data. Eur. J. Epidemiol. 34, 557–565 (2019).

71. Yang, X. et al. Identifying Cancer Patients at Risk for Heart Failure Using Machine Learning Methods. AMIA. Annu. Symp. Proc. 2019, 933–941 (2020).

72. Denny, J. C. et al. PheWAS: demonstrating the feasibility of a phenome-wide scan to discover gene–disease associations. Bioinformatics 26, 1205–1210 (2010).

73. Wu, P. et al. Mapping ICD-10 and ICD-10-CM Codes to Phecodes: Workflow Development and Initial Evaluation. JMIR Med. Inform. 7, e14325 (2019).

74. Denny, J. C. et al. Systematic comparison of phenome-wide association study of electronic medical record data and genome-wide association study data. Nat. Biotechnol. 31, 1102–1111 (2013).

75. Zeng, K., Bodenreider, O., Kilbourne, J. & Nelson, S. J. RxNav: A Web Service for Standard Drug Information. AMIA. Annu. Symp. Proc. 2006, 1156 (2006).

76. American Board of Internal Medicine. ABIM laboratory test reference ranges. January 2024. https://www.abim.org/laboratory-test-reference-ranges.

77. Wang, S., Deng, Z., Chung, F. & Hu, W. From Gaussian kernel density estimation to kernel methods. Int. J. Mach. Learn. Cybern. 4, 119–137 (2013).

78. INSIGHT Clinical Research Network. https://insightcrn.org/. Accessed 30 Oct. 2024.

79. The National Patient-Centered Clinical Research Network. https://www.pcornet.org/. Accessed 2 Feb 2025.

